# The role of wastewater-based epidemiology for SARS-CoV-2 in developing countries: cumulative evidence from South Africa supports sentinel site surveillance to guide public health decision-making

**DOI:** 10.1101/2023.02.13.23285226

**Authors:** Chinwe Iwu-Jaja, Nkosenhle Lindo Ndlovu, Said Rachida, Mukhlid Yousif, Setshaba Taukobong, Mokgaetji Macheke, Laurette Mhlanga, Cari van Schalkwyk, Juliet Pulliam, Tom Moultrie, Wouter le Roux, Lisa Schaefer, Gina Pockock, Leanne Coetzee, Janet Mans, Faizal Bux, Leanne Pillay, Dariah de Villiers, AP du Toit, Don Jambo, Annancietar Gomba, Shaun Groenink, Neil Madgewick, Martie van der Walt, Awelani Mutshembele, Natascha Berkowitz, Melinda Suchard, Kerrigan McCarthy, the SACCESS network

## Abstract

**Background:** The World Health Organisation recommends wastewater based epidemiology (WBE) for SARS-CoV-2 as a complementary tool for monitoring population-level epidemiological features of the COVID-19 pandemic. Yet, uptake of WBE in low-to-middle income countries (LMIC) is low. We report on findings from SARS-CoV-2 WBE surveillance network in South Africa, and make recommendations regarding implementation of WBE in LMICs

**Methods:** Seven laboratories using different test methodology, quantified influent wastewater collected from 87 wastewater treatment plants (WWTPs) located in all nine South African provinces for SARS-CoV-2 from 01 June 2021 – 31 May 2022 inclusive, during the 3rd and 4th waves of COVID-19. Regression analysis with district laboratory-confirmed SARS-CoV-2 case loads, controlling for district, size of plant and testing frequency was determined. The sensitivity and specificity of ‘rules’ based on WBE data to predict an epidemic wave based on SARS-CoV-2 wastewater levels were determined.

**Findings:** Among 2158 wastewater samples, 543/648 (85%) samples taken during a wave tested positive for SARS-CoV-2 compared with 842 positive tests from 1512 (55%) samples taken during the interwave period. Overall, the regression-co-efficient was 0,66 (95% confidence interval=0,6-0,72, R^2^=0.59), but ranged from 0.14-0.87 by testing laboratory. Early warning of the 4^th^ wave of SARS-CoV-2 in Gauteng Province in November-December 2021 was demonstrated. A 50% increase in log-copies SARS-CoV-2 compared with a rolling mean over the previous 5 weeks was the most sensitive predictive rule (58%) to predict a new wave.

**Interpretation:** Variation in the strength of correlation across testing laboratories, and redundancy of findings across co-located testing plants, suggests that test methodology should be standardised and that surveillance networks may utilise a sentinel site model without compromising the value of WBE findings for public health decision-making. Further research is needed to identify optimal test frequency and the need for normalisation to population size, so as to identify predictive and interpretive rules to support early warning and public health action. Our findings support investment in WBE for SARS-CoV-2 surveillance in low and middle-income countries.

**Research in Context:** *Evidence before this study:* Wastewater-based epidemiology (WBE) has long been used to track community disease burden within communities. This approach has become increasingly popular for monitoring the SARS-CoV-2 virus since the beginning of the COVID-19 pandemic. We searched PubMed up until May 2022 using these keywords “SARS-CoV-2”, “COVID”, “wastewater-based epidemiology”, “WBE”, combining them with relevant Boolean operators. We found that majority studies were mostly conducted in high income settings. Huge gap exists for such studies in low and middle income countries, particularly, sub-Saharan Africa. Furthermore, given that WBE of COVID-19 is still in its early stages, more studies are required not only quantify SARS-CoV-2 RNA in wastewater but to also assess the relationship between SARS-CoV-2 in wastewater and clinical case load. Such studies are required to showcase the usefulness of WBE, strengthen the surveillance of COVID-19 and also to improve uptake of these findings by public health officials for decision making.

*Added value of this study:* This is the first study to test a large number of (87) wastewater treatment plants across major cities on a national scale in an African country. Our study not only demonstrates the added value of wastewater-based epidemiology as a great surveillance tool to aid disease control in our setting and similar settings, but it also demonstrates the feasibility of this type of testing. Our research findings are critical for policymakers in South Africa and other low and middle-income countries.

*Implications of all the available evidence:* This study shows that indeed wastewater surveillance can be used to assess the level of disease burden within populations in developing country, especially where there are little or no clinical testing which in turn can inform prompt public health decision. This finding also implies that other infectious diseases which disproportionately affect many low and middle income countries can be monitored using the same approach.

## Background

The World Health Organisation (WHO) recommends wastewater based epidemiology (WBE) for SARS-CoV-2 as a complementary tool for monitoring population-level epidemiological features of the COVID-19 pandemic including the presence or absence of infection, changing trends over time and variant monitoring.[1] Wastewater based epidemiology provides certain advantages over case-based surveillance.[2] These include the ability to monitor large numbers of persons by testing single samples and the potential to estimate the burden of disease in that community from quantitative data.[3–5] WBE eliminates inherent biases in disease burden estimation arising through differential health seeking by infected persons, variable access to clinical laboratory testing and variation in health care practitioner’s propensity to test.[1] WBE may provide early warning of increases in case-incidence [6, 7] enabling prompt public health action to be taken to prevent outbreaks and their consequences.[5] Furthermore, WBE is inherently cost-saving relative to patient-based surveillance systems. Most likely, as a consequence of these advantages, many countries are implementing WBE for SARS-CoV-2. Only one year into the pandemic, a scoping review of the literature showed that 26 countries had already reported the presence of SARS CoV-2 in wastewater including countries in Europe, Americas, Asia, Oceania, and Africa,[8] and by January 2023, 72 countries regularly publish results of SARS-CoV-2 testing in wastewater on 164 dashboards across the world.[9] It is clear that WBE for SARS-CoV-2 contributes to the epidemiological knowledge base that supports individual and public health decision making regarding SARS-CoV-2 prevention, detection and response.

Despite these benefits, and recommendations from developing countries researchers, few African countries have adopted WBE surveillance. Of 164 SARS-CoV-2 WBE dashboards, only two are found in Africa (both in South Africa).[9] In developing countries, structural challenges such as low number of wastewater treatment facilities, low service levels or non-functionality of plants and absent or limited testing resources stand in the way of WBE implementation.[10] Challenges shared with developed countries such as uncertainty regarding how to integrate WBE findings into existing surveillance programmes [11] and how to interpret WBE results with regard to actionable thresholds,[12] have most likely contributed to reluctance to invest in WBE surveillance programmes.

Against this backdrop, the South African Collaborative COVID-19 Surveillance System (SACCESS) network was established in 2021, with the aim of developing standard test methodology, identifying and addressing challenges in qualitative, quantitative detection and RNA sequencing of SARS-CoV-2 in wastewater’.[13]Sites for testing were identified on pragmatic and logistical considerations, and included testing of influent wastewater from almost all treatment plants in densely populated urban metropolitan areas, and single plants in central cities or towns in rural provinces. The SACCESS network actively monitored SARSCoV-2 in wastewater across the country from the third wave of the pandemic in South Africa, where four distinct waves of COVID-19 pandemic have occurred, peaking in June 2020, December 2020, July 2021, and December 2021 respectively[14].

In this paper, we describe the detection and quantification of SARS-CoV-2 from wastewater treatment plants (WWTP) across larger South African metropolitan areas and in single plants from rural provinces from June 2021 to May 2022 (spanning the third and fourth COVID-19 waves). We further explored the relationship between SARS-CoV-2 levels, and district laboratory-confirmed COVID-19 case load, while adjusting for laboratory test method, geographical location, WWTP plant size, testing frequency and timing of testing during or between SARS-CoV-2 waves. We determined the ability of SARS-CoV-2 levels in wastewater to predict a new wave of COVID-19. We reflect on lessons learned to strengthen implementation and design of WBE surveillance networks in middle and low-income countries.

## Methods

### Development of the SACCESS network and extent of surveillance

Commencing in 2018, the NICD tests wastewater for poliovirus as part of the National Department of Health’s polio surveillance programme in accordance with WHO guidance.[15] In 2020, the NICD, and a number of other research, academic and private laboratories commenced with independent projects involving qualitative and quantitative detection of SARS-CoV-2 in local WWTPs. A proof of concept was published by the SAMRC in 2021.[16]

In August 2020, laboratories doing this work collectively established the SACCESS network to support communication of challenges and sharing best practice.[13] Through funding and partnership with the Water Research Commission (WRC), the NICD and partners identified WWTPs for sampling and SARS-CoV-2 testing to maximise coverage across metros and sentinel sites in provinces with smaller populations. A number of webinars were held to share methodologies for concentration, extraction and PCR detection of SARS-CoV-2. [17, 18] Testing of WWTPs commenced between June 2020 and March 2021 as funding became available.

Overall, 87 wastewater treatment sites across nine provinces in SA were tested by the SACCESS network in 2021 (Table S1). This included 40 treatment sites in Gauteng, 5 in Eastern Cape, 5 in Western Cape, 9 in Free State, 12 in KwaZulu-Natal,2 in Limpopo, 3 in Mpumalanga, 2 in Northern Cape province, and 3 in Northwest province. WWTPs were assigned to specific laboratories, and these laboratories consistently tested the same plants over time.

### Laboratory test methods

Each of seven participating laboratories collected samples, concentrated and extracted nucleic acid, and amplified and quantified SARS-CoV-2 RNA using the general principles followed by the NICD methodology, which is fully described here, but using their own RNA extraction and PCR detection methods listed in Table 1. This approach was chosen on account of the number of laboratories participating in the network, the number of plants we wished to test, the different SARS-CoV-2 test kits already used by laboratories, and stock shortages during COVID. The NICD sample collection and processing is as follows: Influent grab samples of one litre volume were collected at treatment facilities during morning peak hours and maintained below 5°C during transportation to the testing facility. No data on flow rate nor volume were collected. At the NICD, 200 mL of sewage sample was centrifuged at 4650 x g (4°C) for 30 minutes. Without disturbing the pellet, 70mL of the supernatant was centrifuged at 3500g for 15 min through the Centricon® Plus-70 centrifugal ultra-filter. The 70mL supernatant was concentrated into a final volume of approximately 1mL. A sewage concentrate volume of 140uL was spiked with 5µL of the commercial internal control (IC) from the Allplex™ 2019-nCoV (Seegene) to monitor RNA inhibitory effects during RNA extraction and RT-qPCR processes. Spiked concentrates plus IC underwent RNA extraction using the QIAmp® viral RNA mini kit (Qiagen) according to the manufacturer’s instructions. Extracted RNA was eluted into 50µL of AVE buffer and tested immediately or stored at -20°C.

**Table 1:**
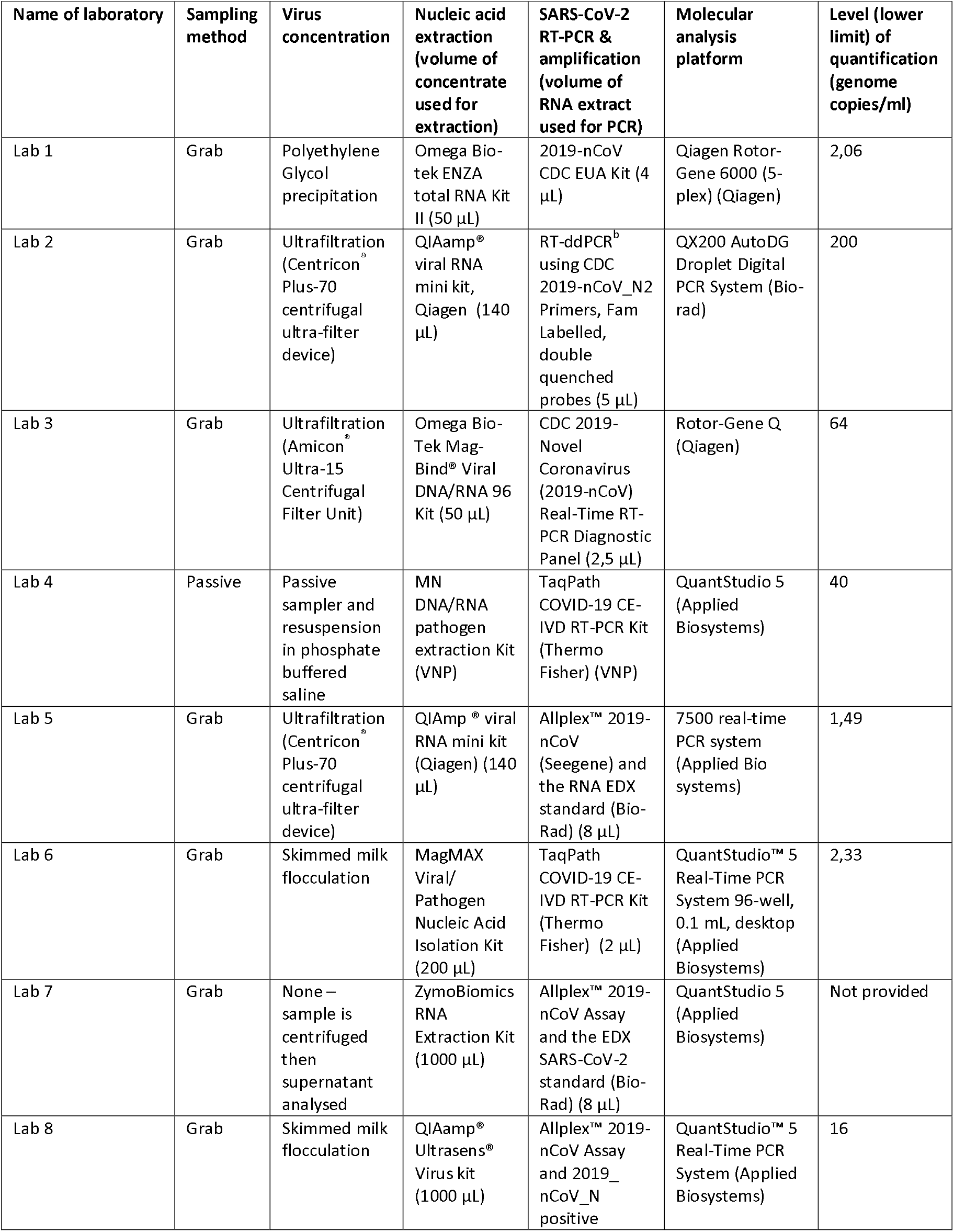
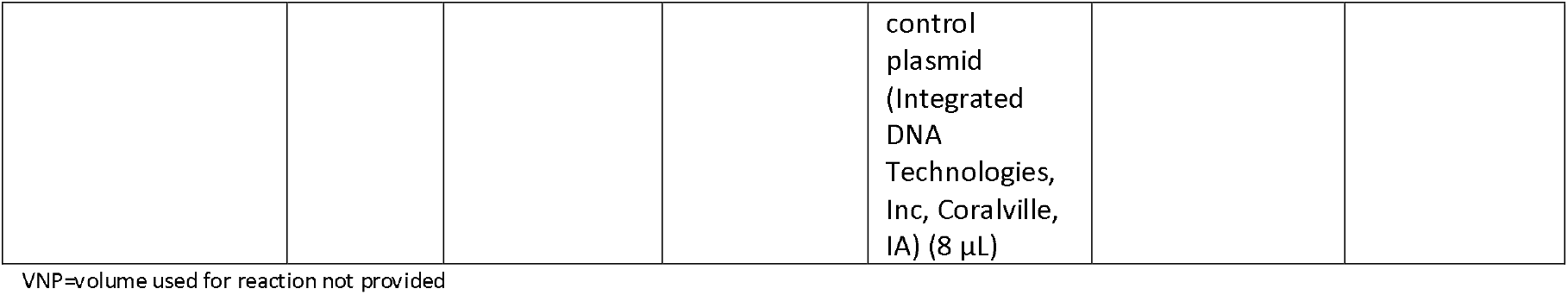
Laboratory methods used for sampling, concentration, RNA extraction volume and method SARS-CoV-2 amplification and detection by partner laboratories.

To prepare for quantification the RNA EDX standard® (Bio-Rad) was double extracted (150 µL and 200 µL) using the QIAmp® viral RNA mini kit (Qiagen) according to manufacturer’s instructions. These extracts were filtered through 1 spin column and eluted into 50µL of AVE buffer. Serial dilutions of the standard were performed at 1:4096, 1:1024, 1:256, 1:64, 1:16, and 1:4. Absolute genome copies for each dilution were calculated as: 2.34, 9.38, 37.50, 150, 600, and 2400 respectively. Four 8.5µL triplicate sets of the standard RNA were prepared for each dilution and stored at -20 °C for simultaneous use with extracted samples.

Finally, viral RNA was detected and quantified in samples using Taqman-based RT-PCR on the 7500 real-time PCR system (Applied Bio systems) using the commercial Allplex™ 2019-nCoV (Seegene) with EDX standard RNA included in each testing run. The PCR reaction mix consisted of 5 µL 2019-nCoV MOM, 5 µL RNase-free H20, 5 µL 5X Real-time One-step Buffer, 2 µL Real-time One-step Enzyme. Cycling conditions were as follows: preheat at 50 °C for 20 min, denature at 95 °C for 15 min, 45 cycles of amplification with 95 °C for 15s and 58 °C for 30s. To increase confidence samples were tested in triplicate, and mean values determined. No participating laboratory normalised for population size.

### Inter-laboratory comparisons and quality assurance

Viral concentrates of 10 samples were randomly selected from the database of each participating laboratory. The concentrates were transported at -20°C to the NICD laboratory where nucleic acid was extracted and SARS-CoV-2 detection was carried out in triplicate. All laboratories achieved the target concordance of 80% with NICD results. In quantification quality assessment panels comprising spiked and unspiked raw wastewater samples and concentrates prepared by the NICD, four of seven laboratories achieved 80% extraction efficiency

### Statistical analysis

We determined the relationship between SARS-CoV-2 case load and SARS-CoV-2 levels in wastewater through correlation and regression analyses as follows. We used the NICD anonymised line list of laboratory-confirmed SARS-CoV-2 cases geo-located to district or sub-district of South Africa. We allocated cumulative case-counts by epi-week and district (or sub-district where available) to corresponding log-transformed weekly SARS-CoV-2 levels (in genome copies per millilitre) from samples obtained from WWTP situated in that district (or sub-district). Where quantification results were below the level of quantification of the test methodology used by each laboratory, we omitted these values from correlation and regression analyses. We conducted correlation and regression analyses for results tested by different laboratories together and independently (Table 2). Regarding correlation, we used Spearman’s correlation test for linear correlations due to the non-normality of data. We conducted simple regression using the case loads of laboratory-confirmed SARS-CoV-2 as the independent variable and multiple regression analyses adjusting for testing laboratory, plant size, frequency of testing (weekly, fortnightly or monthly) and location of plant (by district). We excluded results where wastewater tested negative for SARS-CoV-2 and where the levels of SARS-CoV-2 in genome copies per millilitre were less than the laboratory’s reported level of quantification. For all analyses, a p value <0.05 was considered to be statistically significant. All analyses and graphs were done using Stata (StataCorp, LLC, USA, v 17).

**Table 2.**
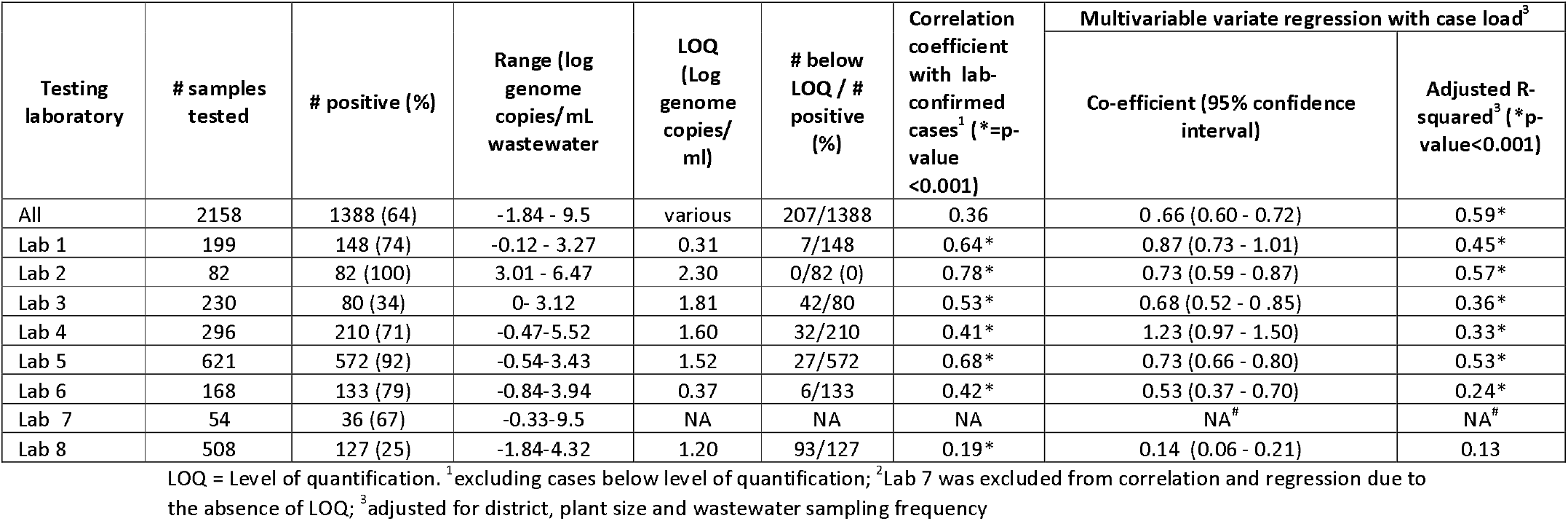
Correlation coefficients and multivariable regression analyses (adjusted for location, size of plant and testing frequency) between SARS-CoV-2 levels (in genome copies/ml) in wastewater and weekly laboratory-confirmed clinical case load of SARS-CoV-2 from 01 June 2021 until 31 May 2022 from 87 plants across all nine South African provinces, for all samples and by testing laboratory.

| Testing laboratory | # samples tested | # positive (%) | Range (log genome copies/mL wastewater) | LOQ (Log genome copies/mL) | # below LOQ / # positive (%) | Correlation coefficient with lab-confirmed cases<sup>1</sup> (*p-value <0.001) | Multivariable variate regression with case load<sup>3</sup> |  |
| --- | --- | --- | --- | --- | --- | --- | --- | --- |
|  |  |  |  |  |  |  | Co-efficient (95% confidence interval) | Adjusted R-squared<sup>3</sup> (*p-value<0.001) |
| All | 2158 | 1388 (64) | -1.84 - 9.5 | various | 207/1388 | 0.36 | 0.66 (0.60 - 0.72) | 0.59* |
| Lab 1 | 199 | 148 (74) | -0.12 - 3.27 | 0.31 | 7/148 | 0.64* | 0.87 (0.73 - 1.01) | 0.45* |
| Lab 2 | 82 | 82 (100) | 3.01 - 6.47 | 2.30 | 0/82 (0) | 0.78* | 0.73 (0.59 - 0.87) | 0.57* |
| Lab 3 | 230 | 80 (34) | 0 - 3.12 | 1.81 | 42/80 | 0.53* | 0.68 (0.52 - 0.85) | 0.36* |
| Lab 4 | 296 | 210 (71) | -0.47-5.52 | 1.60 | 32/210 | 0.41* | 1.23 (0.97 - 1.50) | 0.33* |
| Lab 5 | 621 | 572 (92) | -0.54-3.43 | 1.52 | 27/572 | 0.68* | 0.73 (0.66 - 0.80) | 0.53* |
| Lab 6 | 168 | 133 (79) | -0.84-3.94 | 0.37 | 6/133 | 0.42* | 0.53 (0.37 - 0.70) | 0.24* |
| Lab 7 | 54 | 36 (67) | -0.33-9.5 | NA | NA | NA | NA<sup>#</sup> | NA<sup>#</sup> |
| Lab 8 | 508 | 127 (25) | -1.84-4.32 | 1.20 | 93/127 | 0.19* | 0.14 (0.06 - 0.21) | 0.13 |

LOQ = Level of quantification. <sup>1</sup>excluding cases below level of quantification; <sup>2</sup>Lab 7 was excluded from correlation and regression due to the absence of LOQ; <sup>3</sup>adjusted for district, plant size and wastewater sampling frequency

To investigate the potential that quantitative changes in SARS-CoV-2 wastewater levels could predict a new wave of infection, we identified wastewater results in log-copies of SARS-CoV-2 from samples obtained from 13 WWTP tested by laboratory 1 only. We selected laboratory 1 only to eliminate the impact of variation in test methodology on genome copies/ml and because it had representative sites across South Africa. We paired these by epidemiological week with the positivity rates of SARS-CoV-2 tests performed on persons presenting for COVID-19 investigations in the health districts where these plants are located. We calculated the sensitivity and specificity of a ‘wastewater test’ to detect a new wave as defined by the ‘gold-standard’, which we defined as a positivity rate amongst clinical PCR tests exceeding 10% on the upward trajectory of a wave. We chose to utilise the clinical positivity rate to define our gold standard, rather than the National Department of Health’s (NDoH) definition of a wave based on incidence rates [14], as positivity rate may more sensitively detect a rise in population burden of disease and is more robust in changes to testing behaviour. We defined the wastewater ‘test’ as a Y% increase in log-gene copies in the week of interest, compared to the average of the last Z weeks, where Z = 1 to 6 weeks and Y varied from 5% to 200%. In the main analyses of predictive indicators, we calculated the sensitivity of the ‘test’ as the fraction of times our ‘wastewater test’ in the epidemiological week of interest, detected the crossing of the 10% threshold X= 1-, 2-, 3- or 4-weeks’ ahead. In supplementary analyses (see supplementary material), we also investigated tests based on any of the N previous weeks. The downward trajectory of a wave defines the ‘gold-standard’ for the specificity calculation. Regarding specificity, our ‘test’ should not predict a wave of new infection between the time points that positivity drops below 10% for the first time and the lowest positivity rate between two waves. Sensitivity calculations were based on as many available data points as possible (between 9 and 22 data points). Each of the 15 sites contributed at most one data point for each of two waves. Specificity calculations were based on between 103 and 123 data points, since more than one-time point in the downward trajectory was considered. However, data was missing during some weeks, which reduced the number of data points available for calculations. These methods are further described in the Supplemental Methods for details.

## Results

A total of 2,158 wastewater samples were tested by the SACCESS network between 01 June 2021 – 31 May 2022 inclusive, coinciding with the 3rd and 4th waves of the COVID-19 pandemic. Of these, 1388 (64%) tested positive for SARS-CoV-2. During the 3^rd^ and 4^th^ waves, 638 tests were conducted and 543 (85%) were positive. During the interwave periods, 1,512 samples were tested and of these, 842 (55%) were positive. Amongst all positive tests 207/1,388 were below the limit of quantification of the test method used by the testing laboratory and were excluded from correlation and regression analyses.

Figures 1A-F show the times series of laboratory confirmed cases by district and log genome copies of SARS-CoV-2/mL of wastewater-for WWTPs across all study metros for the entire review period, excluding those WWTPs with three or fewer data points after elimination of results below the level of quantification. On visual inspection of time series graphs, high concentrations were mostly seen during the waves of the pandemic, and low concentrations during the interwave period. The levels of SARS-CoV-2 from sentinel plants in Gauteng subdistricts that were tested by the NICD from epidemiological week 40 (just before the onset of fourth wave) are illustrated in Figure 2 and demonstrate a rise in levels which precede the increase in cases.

**Figure 1 A-F.**
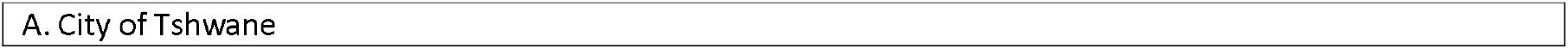
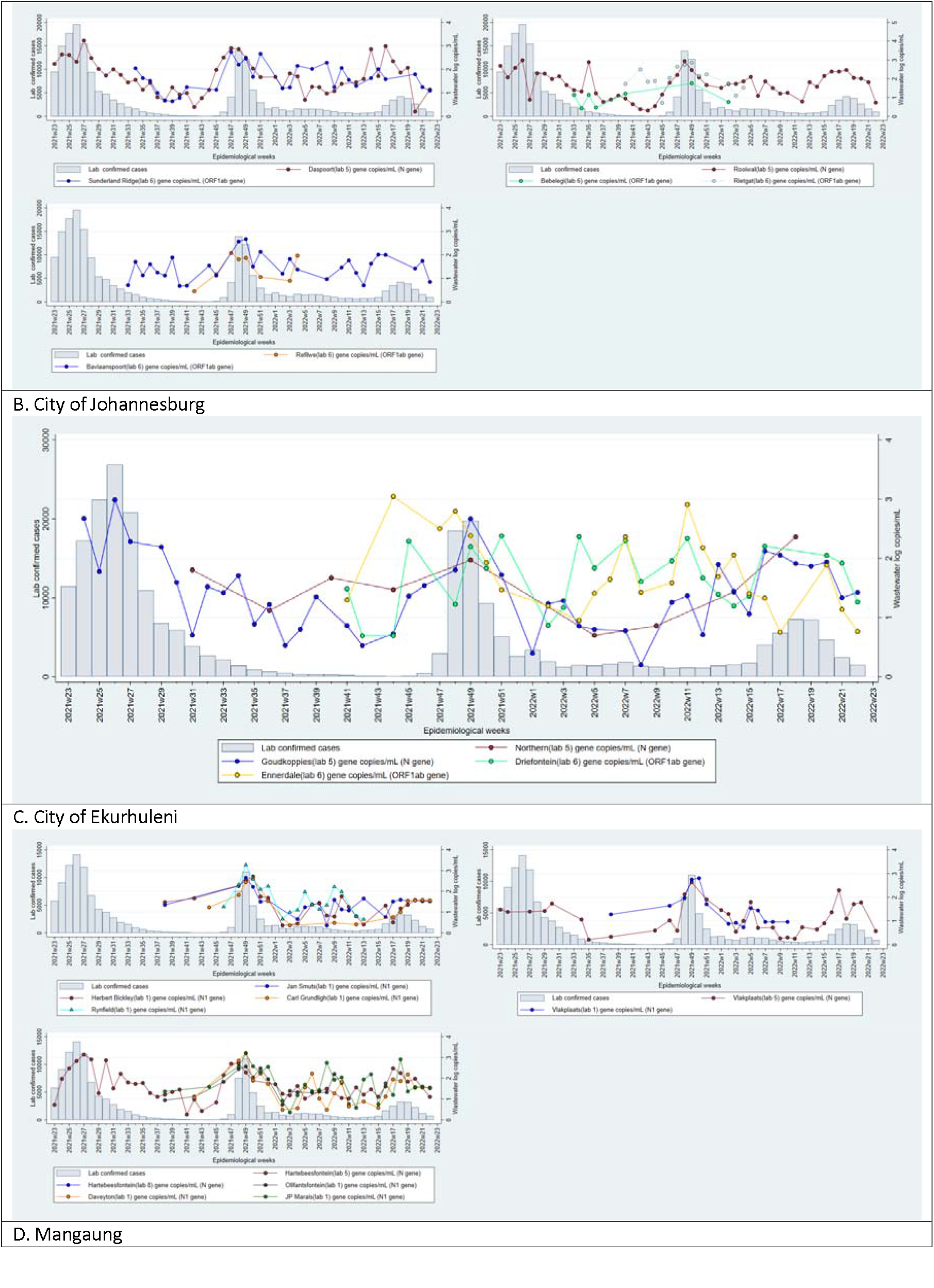
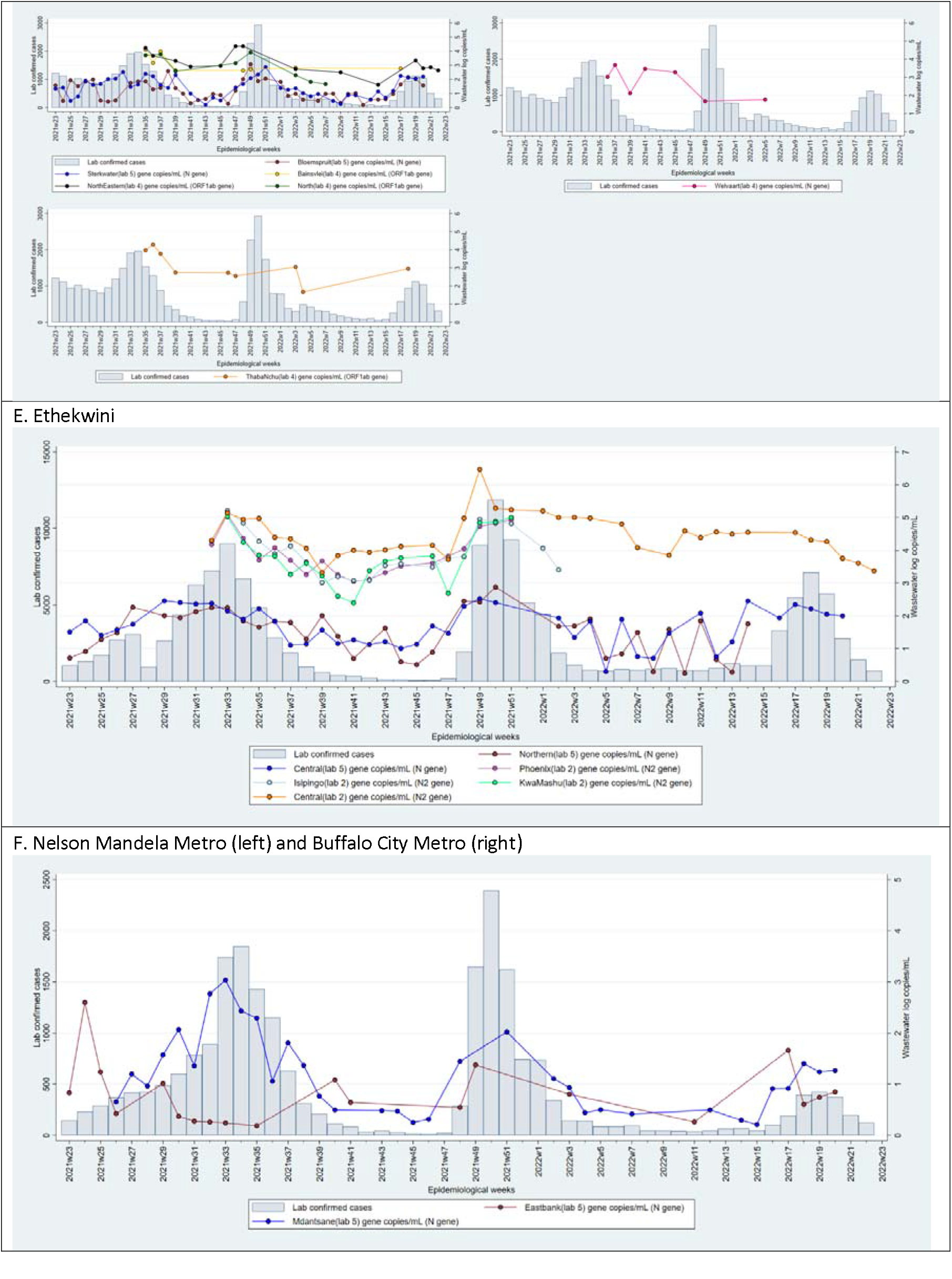
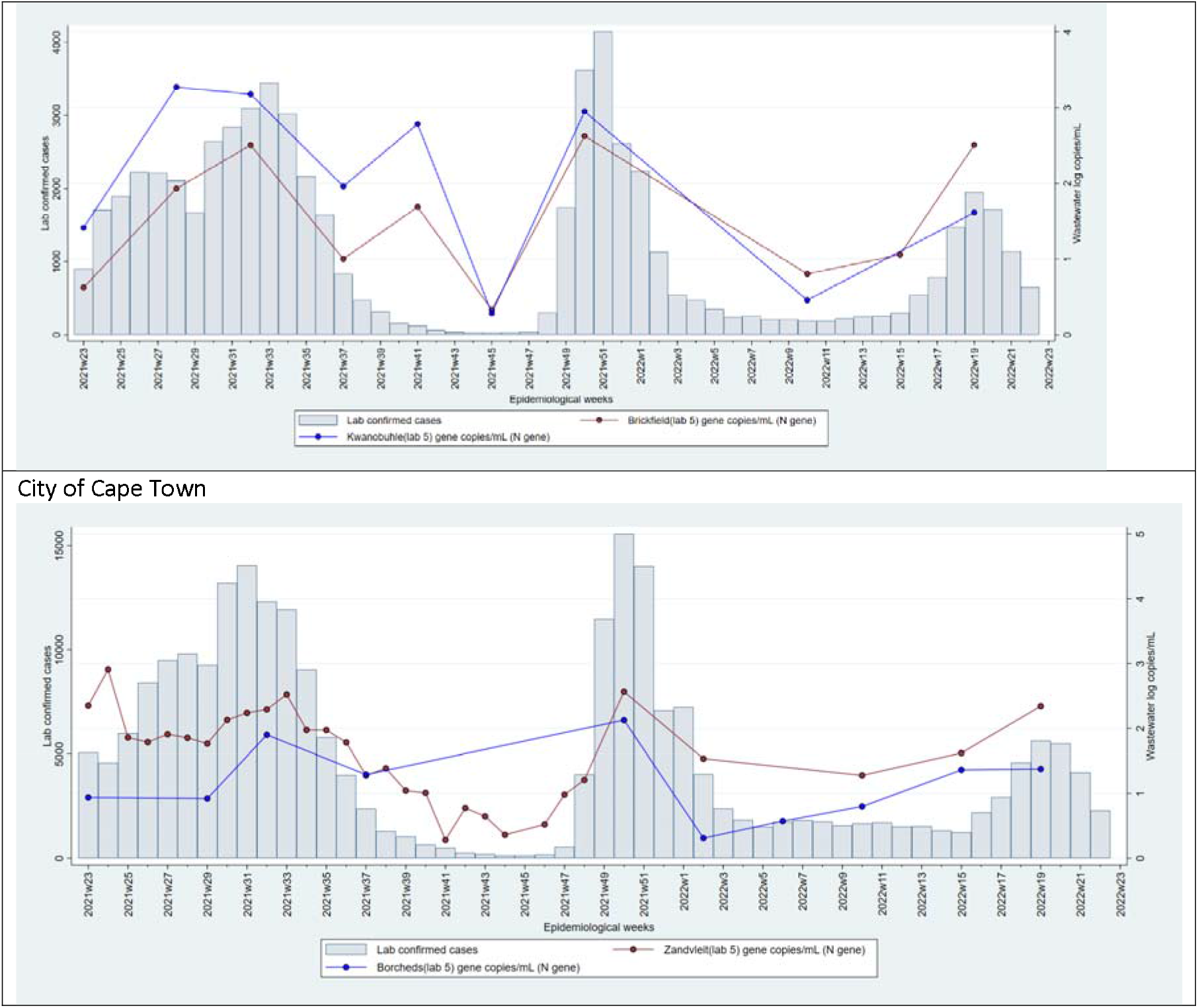
Laboratory-confirmed cases (left axis, grey bars) and log genome copies SARS-CoV-2 per millilitre of wastewater (right axis, lines), by epidemiological week. Colors represent different wastewater testing plants. Results below the level of quantification of each laboratory are omitted, as are wastewater treatment plants with 3 or fewer results. The testing laboratory (1-7) is indicated in the key.

**Figure 2.**
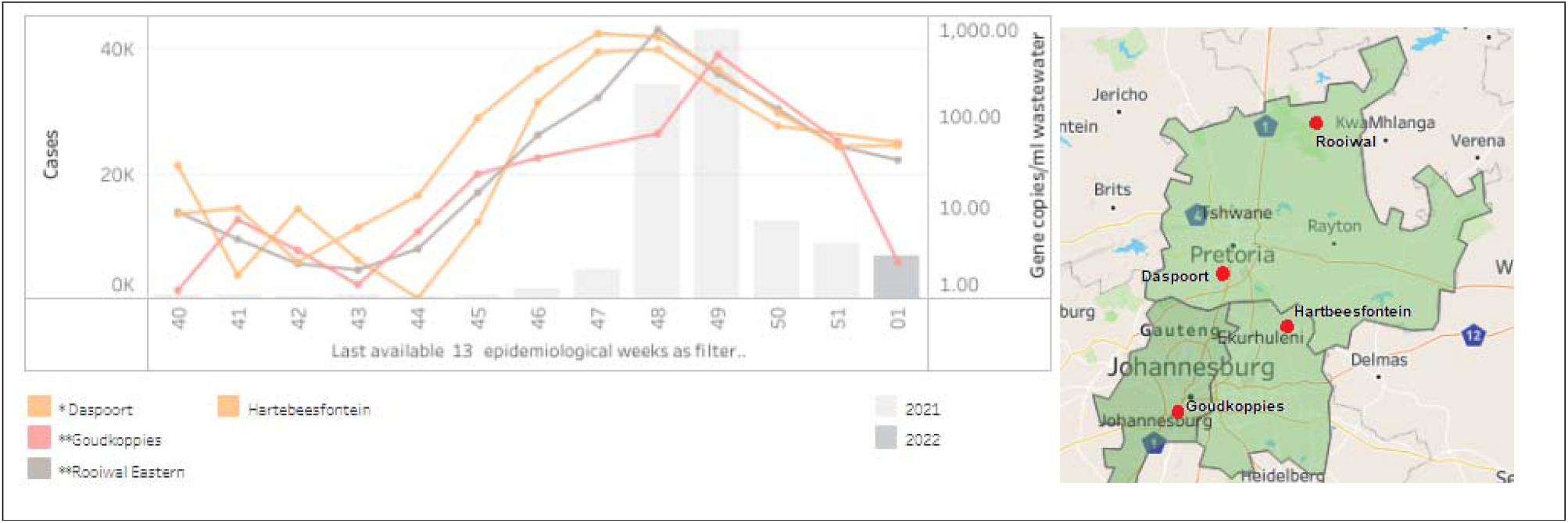
SARS-CoV-2 levels in wastewater (genome copies/ml) for four wastewater treatment plants in Gauteng (see inset map) and case load of laboratory-confirmed SARS-CoV-2 cases in Gauteng Province by epidemiological week 2021

Table 2 lists the correlation and multivariable regression coefficients (adjusted for location, size of plant and testing frequency) between SARS-CoV-2 levels (in genome copies/ml) and weekly laboratory-confirmed case loads in the corresponding district/sub-district by testing laboratory. Table 3 presents results of the identical analyses but conducted for plants located in Gauteng Province only, where the clinical epidemiology of SARS-CoV-2 and case loads to which wastewater levels of SARS-CoV-2 were compared were similar by district in Gauteng. Correlation co-efficient between SARS-CoV-2 genome copies/ml wastewater and laboratory-confirmed cases by district for each specific plant are shown in the Supplement Table S1.

**Table 3.**
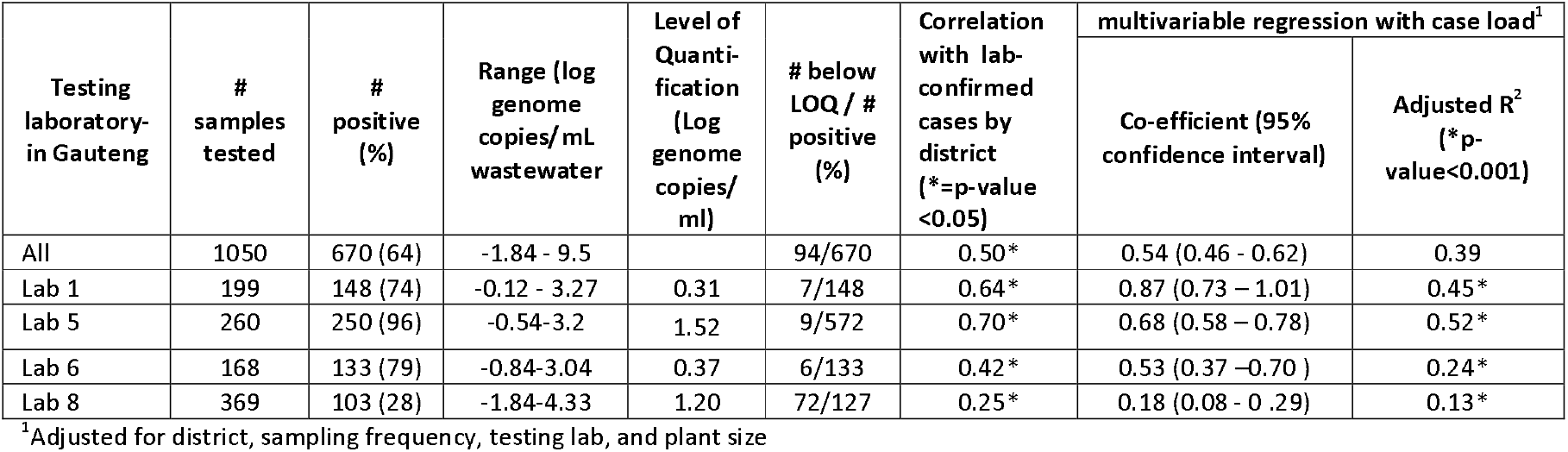
Correlation coefficients and multivariable regression analyses (adjusted for location, size of plant and testing frequency) between SARS-CoV-2 levels (in genome copies/ml) in wastewater and weekly laboratory-confirmed clinical case load of SARS-CoV-2 from 44 wastewater treatment plants in Gauteng Province from 01 June 2021 until 31 May 2022.

There was a significant association between wastewater levels and case load in the multivariate linear regression after adjusting for testing laboratory, district, sampling frequency, and plantsize (p<0.001, R2 =0.6). Similarly, a significant association was seen when the sample regression model was restricted to the Gauteng province where the clinical epidemiology of SARS-CoV-2 and case load to which wastewater levels of SARS-CoV-2 were compared were similar by district in Gauteng (p<0.001, R2 =0.4).

Regarding a predictive rule, sensitivity was highest for the lead period of one and two weeks, and when the average log genome copies/ml included values up to six preceding weeks from the point of measurement. The time points at which the rule was generated that were further from the point of declaration of a wave (a clinical positivity rate of > 10%) were generally less sensitive. None of the tests with specified lead times had sufficiently high sensitivity and specificity to be provide robust indicators of the start of a new wave (Figure 3). The most informative indicator we identified was whether or not the wastewater level in any of the previous 5 weeks was at least 100% (2X) higher than the average of the previous 5 weeks; this indicator had a sensitivity of 79% and a specificity of 78% (Figure S1).

**Figure 3:**
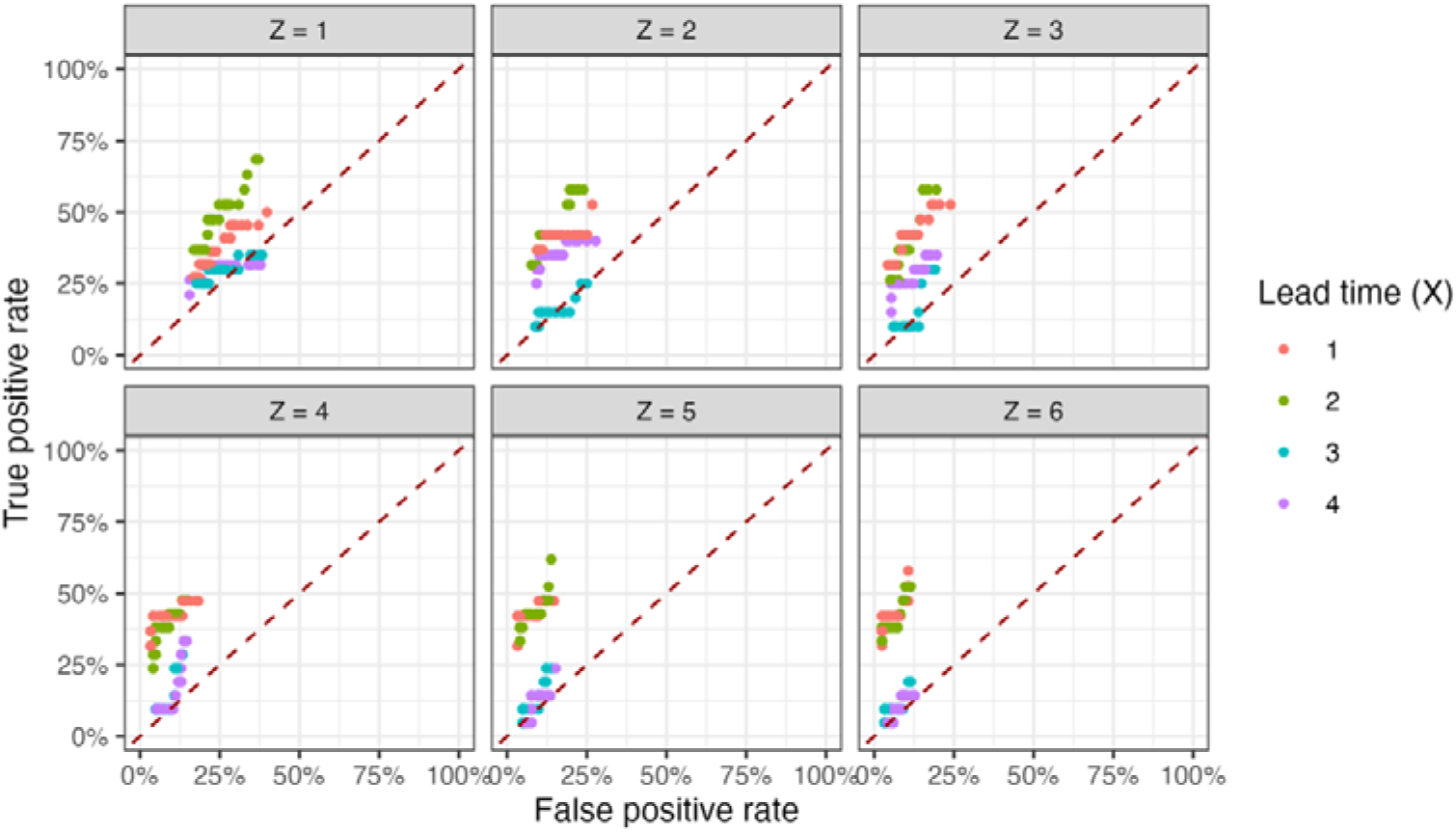
True positive rate (sensitivity) and false positive rate (1-specificity) measures of a defined rule vs gold standard to establish the ability of wastewater levels of SARS-CoV-2 to predict an epidemic wave. The ‘gold-standard’ for sensitivity is defined by the epidemiological week when the positivity rate of clinical testing exceeds 10% on the upward trajectory of a wave. The ‘gold-standard’ for specificity is defined on the downward trajectory of a wave, at the time point when the positivity rate drops below 10% for the first time and the lowest positivity rate between two waves. The test used in this figure is defined as whether or not the SARS-CoV-2 log-gene copies in wastewater X weeks ago exceeded the average log-gene copies of the previous Z weeks by a percentage varied from 5% to 200%.

## Discussion

Results of quantitative SARS-CoV-2 testing across South African wastewater treatment plants have demonstrated good correlation with district laboratory-confirmed COVID-19 case load, and provided early warning of the 4^th^ wave of SARS-CoV-2 in Gauteng Province in November-December 2021. Our results demonstrated variation in the strength of correlation across testing laboratories, and redundancy of findings across co-located testing plants, suggesting that test methodology should be standardised and that surveillance networks may utilise a sentinel site model without compromising the value of WBE findings for public health decision-making. Further research is needed to identify optimal test frequency so as to identify predictive and interpretive rules to support early warning and public health action. Our findings support investment in WBE for SARS-CoV-2 surveillance in low and middle-income countries.

Our data series, comprising 1388 observations over 2 waves of COVID-19, illustrate that the levels of SARS-CoV-2 in wastewater follow trends in laboratory-confirmed SARS-CoV-2 case load, and demonstrate high correlation and strong regression analysis indicators between case load and genome copies/ml from single WWTPs over time (Supplementary table). These findings concur with global observations regarding the usefulness of WBE of SARS-CoV-2 as an indirect measure of population burden of disease [7] and support the recommendations of the WHO interim guidance on Environmental surveillance for SARS-CoV-2, that WBE provides insight into the distribution and time trends in SARS-CoV-2 levels at community level.[1] Our findings, generated at a time when SARS-CoV-2 clinical testing was widespread and accessible, point to the usefulness of WBE to provide insight into the distribution of cases and relative burden of disease as clinical testing rates decline and hospitalisation rates decrease with subsequent waves of infection.[19]

The variation in the strength of correlation between wastewater levels of SARS-CoV-2 and laboratory-confirmed case load between plants tested by different laboratories, even when plants were co-located in the same health district where timing of the wave and burden of clinical disease to which the wastewater levels were compared, were constant. It is important to understand and address variation in SARS-CoV-2 levels, as variation may contribute to weaker correlation with clinical burden of disease and may therefore undermine the potential usefulness of WBE data.

A key source of variation in our context arose from the different sample collection, extraction and detection methods, and different wastewater sample volumes used by participating laboratories to determine quantitative SARS-CoV-2 levels. Passive sampling, as utilised by one of our laboratories, will trap particulate matter, and will report consistently higher wastewater levels compared with grab samples, the method preferred by most of our laboratories, as RNA seems to concentrate more in solid phase of the wastewater matrix [20, 21]. Differences in concentration and RNA extraction techniques and the volume of concentrate that undergoes RNA extraction will retain more or less template RNA for detection.[22] Finally, the different genes of SARS-CoV-2 that are detected and amplified by different RT-PCR detection kits, may be present at variable amounts in wastewater because of differential decay of viral fragments in a hostile environment.[23] We also observed variable extraction efficiencies in the quality assessment panels (data not shown), suggesting that some methodologies are more sensitive and precise than others, a finding observed elsewhere.[24] Special attention should be given to digital PCR technologies, as we observed higher levels, stronger correlation and closer tracking of clinical cases with results from the laboratory using this technology [24].

Sources of variation in SARS-CoV-2 levels in wastewater other than those that may be ascribed to test methodology [25, 26] may also have accounted for variation in strength of correlation with clinical case load. In our network, these may include the following: Firstly, the district boundaries to which laboratory-confirmed case loads were allocated, do not correspond with sewage reticulation networks. We were unable to reconfigure cases to provide data from populations resident within the sewage catchment areas. If different districts reported different case loads correlation of results from a single plant serving populations across district boundaries may be weaker. Secondly, the WWTPs from where samples were taken were large, serving a median of 100,000 persons (range 4,000-1,200,000). The geographical areas served by these WWTPS were socio-economically diverse. It is well established that high population density and poorer socio-economic conditions are associated with higher SARS-CoV-2 transmission and therefore higher disease burden, hospitalisation and mortality rates.[27] Yet these areas may be under-represented in sewage networks because of informal settlements, or dilution with effluent from more well-resourced areas may artificially lower SARS-CoV-2 levels. Thirdly, the high proportion of asymptomatic disease [28], and differential access to SARS-CoV-2 testing may have led to discontinuity between wastewater levels and clinical cases [29]. Fourthly, and particularly problematic in larger networks, differential SARS-CoV-2 RNA decay rates within the sewage network [27], destruction of RNA through mixing with industrial effluent [19], and dilution of RNA through environmental sources such as rain-water may alter the relationship to clinical case load. Finally, but less likely in large wastewater treatment plants, variation in population size over time through in-and outward migration may artificially change SARS-CoV-2 levels in wastewater. These sources of variation may explain why some testing laboratories, particularly those testing plants in the rural provinces with smaller and less well-resourced communities demonstrated lower correlation with case load. These sources of variation may also explain why our data did not support our attempt to create a highly sensitive and generalizable predictive rule for a new clinical wave.

We chose not to obtain additional contextual data pertaining to wastewater flow rates and volumes, nor did we attempt to normalise data to population load, all of which would have allowed us to estimate burden of disease due to COVID-19 in sewer catchment areas. This is a key differentiating factor in our approach compared with well-resourced contexts. The Netherlands, for example, publishes the number of SARS-CoV-2 particles per 100,000 persons by health district.[30] Our decision was based on a number of factors, most notably that only 60% of South African population is served by waterborne sewage, and a smaller proportion of that served by wastewater treatment networks, the balance using septic tanks.[31] Other factors include the difficulty of standardising PCR methods to support normalisation, unavailability of sewage reticulation network maps and therefore uncertainties regarding precise geographical areas being tested, and uncertainties regarding decay rates and degradation of RNA within complex industrial and residential sewage networks.

Under the current circumstances, and with experience learned from our network, we reflect on the suitability of the current configuration of wastewater testing SARS-CoV-2 for surveillance purposes. The sources of variation (including laboratory test methodology) that lead to differing correlation and regression indicators between levels and case-load, and the relatively high proportion of that is not served by wastewater treatment plants, suggest as we anticipated, that estimation of precise population burden of disease in our context is presently unattainable. Further, testing large numbers of samples from geographically disparate plants creates high workloads and sample transport costs. Regarding SARS-CoV-2 levels, we observed that plants co-located in the same district revealed similar temporal trends, suggesting redundancy in the data to support public health decision-making authorities. It is our opinion therefore, that extensive networks that test influent from large wastewater treatment plants across entire geographical areas may not be useful in low and middle income settings from a public health perspective.

Therefore, given these limitations in our middle-income context, we propose a sentinel site WBE surveillance model with centralised testing to eliminate variation due to different laboratory test methods. Such a network serves public health decision-making needs by providing relative population burden, temporal trends and geographic distribution of viral infection and variants. These data may be triangulated with results from clinical testing to provide a fuller picture of disease epidemiology. In the context of a such a model, with carefully chosen sites that ensure maximal population representativeness (including wastewater treatment plants, environmental sources, rivers downstream of informal settlements) it may be possible with further research, to determine of the population burden of disease. Key research questions include the added value of more frequent testing normalising for population size, measuring wastewater flow-rates, and correcting for environmental factors such as ambient temperature and dilution with rainfall in defined sewage reticulation networks where populations are well characterised. These findings may serve public health decision-making at a local level, and may inform generalisability of results from sentinel surveillance sites. Presently, despite its many advantages and potential, WBE cannot suffice as a stand-alone surveillance methodology in our or any context. Its key limitation is the inability to provide insight into disease severity. Therefore, WBE findings should always be interpreted with other measures of population health, including hospitalisation and mortality data.

## Conclusion

We have presented findings from the wastewater-based surveillance for SARS-CoV-2 during the 3^rd^ and 4^th^ waves of SARS-CoV-2 in South Africa which demonstrate both the value and limitations of wastewater-based epidemiology in low and middle income countries. Further, our work has highlighted key areas for investigation to support interpretability of quantitative SARS-CoV-2 levels in wastewater with reference to burden of disease, and provided evidence to support the optimal configuration of WBE surveillance networks. Given trends in viral evolution and immune evasion, SARS-CoV-2 will remain a concern for population health. Investment in WBE as a surveillance tool will continue to support provision of data to address key uncertainties in decision-making aimed at preserving population health.

## Data Availability

All data produced in the present study are available upon reasonable request to the authors

## Declarations

### Ethics approval and consent to participate

Not applicable

### Data sharing

Data used for this study is available on request

## Authors’ contributions

Study was conceived by KM, MS and the SACCESS network; sample collection, processing, laboratory testing and generation of results were done by NN, SR, MY, WL, LS, GP, LC, JM, FB, LP, DV, AD, DJ, AG, SG, NM, MV, AM, NB; data visualisation and analyses were done by CI, NN, KM, LM, CS, JP and TM; all authors contributed to the writing of various sections of the manuscript; CI, NN and KM compiled the first draft of the manuscript; all authors read and approved the final version of the manuscript

## Acknowledgements

We wish to acknowledge the following in no particular order:

- The NICD Centre for Respiratory Disease and Meningitis are thanked for their assistance in setting up and troubleshooting PCR testing, and ongoing supportive collaboration
- The staff from participating labs
- The contributions of local government and wastewater treatment staff to sample collection and transport is acknowledged and appreciated.
- Students who supported with sample collections and processing the samples: Mr Thoriso Mooa, Ms Unarine Matodzi, Ms Phiwinhlanhla Nkosi – SAMRC-TB Platform
- The Water Research Commission, for their vision and support
- The NICD SARS-CoV-2 epidemiology and IT team members for sharing district and sub-district case burdens in order to generate graphs. These team members include Andronica Moipone Shonhiwa, Genevie Ntshoe, Joy Ebonwu, Lactatia Motsuku, Liliwe Shuping, Mazvita Muchengeti, Jackie Kleynhans, Gillian Hunt, Victor Odhiambo Olago, Husna Ismail, Nevashan Govender, Ann Mathews, Vivien Essel, Veerle Msimang, Tendesayi Kufa-Chakezha, Nkengafac Villyen Motaze, Natalie Mayet, Tebogo Mmaborwa Matjokotja, Mzimasi Neti, Tracy Arendse, Teresa Lamola, Itumeleng Matiea, Darren Muganhiri, Babongile Ndlovu, Khuliso Ravhuhali, Emelda Ramutshila, Salaminah Mhlanga, Akhona Mzoneli, Nimesh Naran, Trisha Whitbread, Mpho Moeti, Chidozie Iwu, Eva Mathatha, Fhatuwani Gavhi, Masingita Makamu, Matimba Makhubele, Simbulele Mdleleni, Tsumbedzo Mukange, Trevor Bell, Lincoln Darwin, Fazil McKenna, Ndivhuwo Munava, Muzammil Raza Bano, Themba Ngobeni
- The Staff of SACCESS network laboratories, for their assistance in generating these results

