## Supplementary material for "The role of wastewater-based epidemiology for SARS-CoV-2 in developing countries: cumulative evidence from South Africa supports sentinel site surveillance to guide public health decision-making"

### Supplementary methods: description of the positivity data

The estimates of weekly proportions testing positive at a municipal district level (and for metros) used in the study are derived from the data extracted from the database of Covid tests maintained by the National Institute for Communicable Diseases. These data form the backbone of the weekly testing reports issued by the NICD (<https://www.nicd.ac.za/diseases-a-z-index/disease-index-covid-19/surveillance-reports/weekly-testing-summary/>).

The data used in this study reflect the 20.82 million tests conducted by public and private service providers and laboratories from March 2020 through to early March 2022 that are captured on the NICD database.

Since lags occasionally present in the reporting of results to the NICD (and their subsequent inclusion in the database), the temporal aspect of the data is determined by the date of specimen collection – not the date of reporting of the results to the NICD. These were then aggregated into the epi-weeks used by the NICD (weeks running from a Sunday to the subsequent Saturday).

The unit-record data on each test do not contain geolocatable addresses of the subjects being tested. To analyse the testing data geospatially, the name of the facility where the specimen was collected is used as a proxy for residence. Two datasets, giving the geolocation (latitude and longitude, typically to 4-6 decimal places) of almost every public facility, and the vast majority of private facilities, in the country were constructed. While the proportion of tests that were able to be geolocated in this fashion varies from week to week and with incremental augmentations to the datasets, the procedure is routinely able to geolocate around 98-99% of weekly tests conducted in public facilities and between 78 and 85% of weekly tests conducted in private facilities.

Using standard GIS ray-casting algorithms, each test with a geolocatable facility name was able to be allocated to a municipal district (and health district within the country’s metropolitan areas).

Finally, to eliminate differentials in the age-sex profile of those testing across municipalities in a given week and to facilitate drawing of comparisons across districts, the estimate of the district-specific proportions testing positive were standardised by fitting a logistic regression model to the tests in a given week, and then – using a margins-at-mean approach – deriving the estimate of the age-sex standardised proportion testing positive in a particular district on the assumption that age-sex profile testing in that week in any district was that of the overall age-sex profile testing in that week.

The results of these manipulations is an analytical dataset, by week and district, of the resulting standardised proportions testing positive and the number of tests conducted. (It must, however, be appreciated that the number of cases in any week in any district cannot be directly inferred by multiplying the number of tests by the standardised proportions testing positive).

### Supplementary Figure

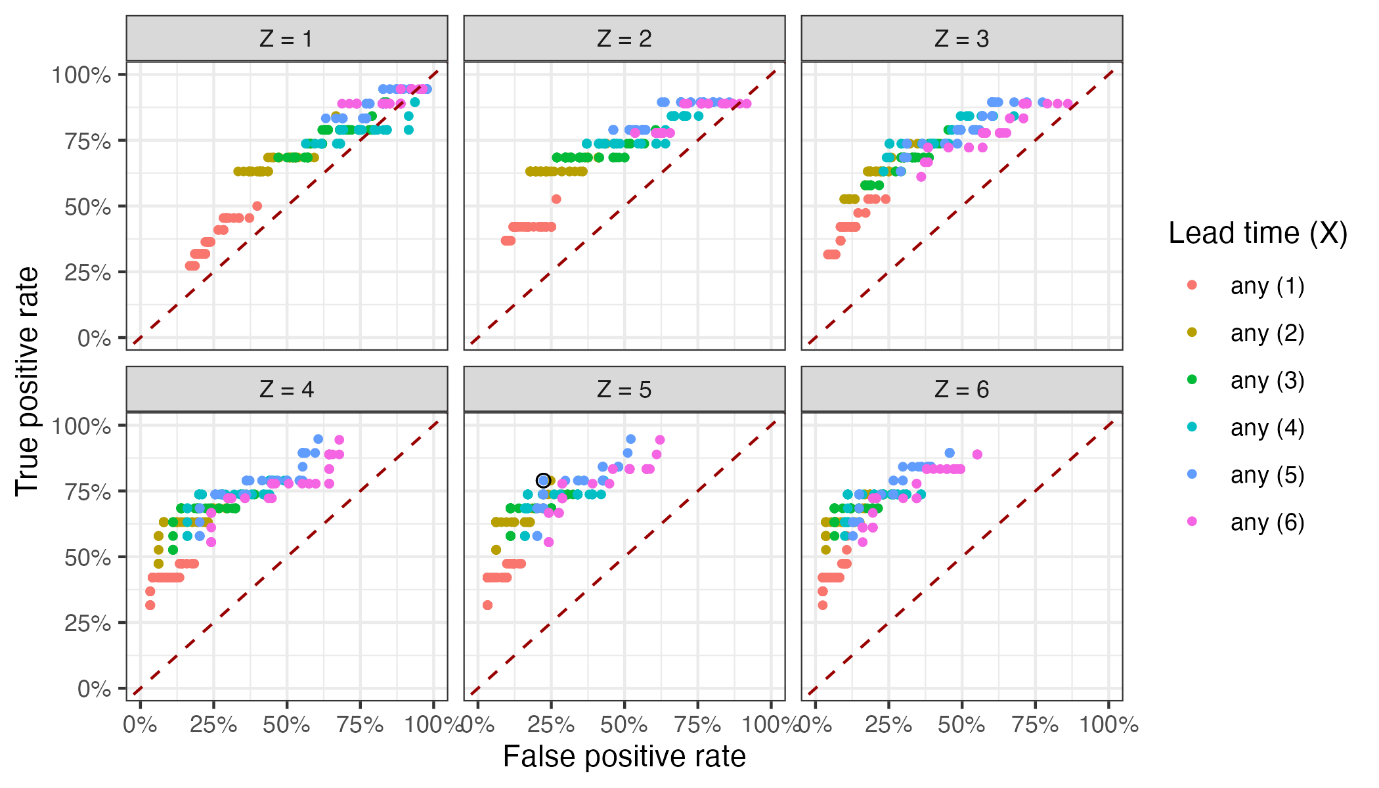

Figure S1. True positive rate (sensitivity) and false positive rate (1-specificity) measures of a defined rule vs gold standard to establish the ability of wastewater levels of SARS-CoV-2 to predict an epidemic wave. The ‘gold-standard’ for sensitivity is defined by the epidemiological week when the positivity rate of clinical testing exceeds 10% on the upward trajectory of a wave. The ‘gold-standard’ for specificity is defined on the downward trajectory of a wave, at the time point when the positivity rate drops below 10% for the first time and the lowest positivity rate between two waves. The test used in this figure is defined as whether or not any of the SARS-CoV-2 log-gene copies in wastewater in any of the past X weeks exceeded the average log-gene copies of the previous Z weeks by a percentage varied from 5% to 200%.

### Table S1: Characteristics of wastewater testing plants and samples tested

| **Wastewater treatment plant by testing laboratory** | **Province** | **District** | **Plant size** | **Plant size category** | **Sampling frequency** | **No of samples tested (# positive for SARS-CoV-2)** | **Correlation coefficient (p<0.005)** | **Regression** | | |
| --- | --- | --- | --- | --- | --- | --- | --- | --- | --- | --- |
|  |  |  |  |  |  |  |  | **Adjusted R2**  **(p<0.005)** | **Co-efficient** | **95% CI** |
| **Lab 1** | **Province 1** |  |  |  |  |  |  |  |  |  |
| Lab1_WWTP1 | Province 1 | Prov1_district 1 | 10000 | Small | Fortnightly | 22 (17) | 0.8 | 0.7 | 1.3 | 0.88 - 1.68 |
| Lab1_WWTP2 | Province 1 | Prov1_district 1 | 100000 | Medium | Fortnightly | 28 (16) | 0.5 | 0.3 | 0.80 | 0.33 - 1.26 |
| Lab1_WWTP3 | Province 1 | Prov1_district 1 | 30000 | Medium | Fortnightly | 27 (22) | 0.7 | 0.5 | 0.97 | 0.61 -1.33 |
| Lab1_WWTP4 | Province 1 | Prov1_district 1 | 100000 | Medium | Fortnightly | 28 (24) | 0.4 # | 0.26 | 0.75 | 0.30 - 1.20 |
| Lab1_WWTP5 | Province 1 | Prov1_district 1 | 50000 | Medium | Fortnightly | 28 (24) | 0.6# | 0.40 | 0.99 | 0.52 - 1.45 |
| Lab1_WWTP6 | Province 1 | Prov1_district 1 | 200000 | Large | Monthly | 22 (15) | 0.8 | 0.64 | 0.86 | 0.64 - 1.08 |
| Lab1_WWTP7 | Province 1 | Prov1_district 1 | 100000 | Medium | Fortnightly | 6 (4) | 0.8 | -0.01 | 1.00# | -3.34- 5.35 |
| Lab1_WWTP8 | Province 1 | Prov1_district 1 | 350000 | Large | Weekly | 22 (16) | 1.0 | 0.74# | 0.67# | -2.62 - 3.96 |
| **Total** |  |  |  |  |  | **183 (138)** |  |  |  |  |
| **Lab 2** |  |  |  |  |  |  |  |  |  |  |
| Lab2_WWTP1 | Province 2 | Prov2_district 1 | 73487 | Medium | Weekly | 20 (20) | 0.8 | 0.63 | 0.75 | 0.49 - 1.02 |
| Lab2_WWTP2 | Province 2 | Prov2_district 1 | 350000 | Large | Weekly | 21 (21) | 0.7 | 0.38 | 0.67 | 0.42 – 0.92 |
| Lab2_WWTP3 | Province 2 | Prov2_district 1 | 266744 | Large | Weekly | 19 (18) | 0.8 | 0.64 | 0.75 | 0.49 - 1.02 |
| Lab2_WWTP4 | Province 2 | Prov2_district 1 | 108666 | Medium | Weekly | 18 (18) | 0.8 | 0.63 | 0.68 | 0.42 - 0 .94 |
| **Total** |  |  |  |  |  | **78 (76)** |  |  |  |  |
| **Lab 3 GreenHill/Praecautio** | Province 2 | Prov2_district 1 | 1500 | Small |  |  |  |  |  |  |
| Lab3_WWTP1 | Province 2 | Prov2_district 1 | 37564 | Medium | Fortnightly | 6 (2) | ## | ## | ## | ## |
| Lab3_WWTP2 | Province 2 | Prov2_district 1 | 4494 | Small | Fortnightly | 18 (3) | 0.4# | 0.05 | 0.46# | -0.33 - 1.25 |
| Lab3_WWTP3 | Province 2 | Prov2_district 1 | 71771 | Medium | Fortnightly | 18 (10) | 0.3# | 0.05 | 0.57# | -0.38 - 1.53 |
| Lab3_WWTP4 | Province 2 | Prov2_district 1 | 5142 | Small | Fortnightly | 11 (7) | 0.6# | 0.40 | 1.05# | 0.14 - 1.97 |
| Lab3_WWTP5 | Province 2 | Prov2_district 2 | 5142 | Small | Fortnightly | 1 (1) | 0.7# | 0.40 | 1.30# | 0.38 - 2.23 |
| Lab3_WWTP6 | Province 2 | Prov2_district 2 | ** | ** | Fortnightly | 29 (16) | 0.7 | 0.44 | 0.94 | 0.36 - 1.53 |
| Lab3_WWTP7 | Province 2 | Prov2_district 1 | 21639 | Medium | Fortnightly | 28 (6) | 0.5# | 0.17 | 0.91# | -0.17 – 2.00 |
| Lab3_WWTP8 | Province 3 | Prov3_district 1 | 43100 | Medium | Monthly | 32 (13) | 0.5# | 0.46 | 0.91 | 0.44 - 1.37 |
| Lab3_WWTP9 | Province 3 | Prov3_district 1 | 30400 | Medium | Fortnightly | 21 (10) | 0.45# | 0.43 | 0.70# | 0.23 - 1.17 |
| Lab3_WWTP10 | Province 3 | Prov3_district 1 | 48700 | Medium | Fortnightly | 26 (15) | 0.5# | 0.53 | 0.86# | 0.30 - 1.41 |
| Lab3_WWTP11 | Province 3 | Prov3_district 1 | 25400 | Medium | Weekly | 18 (7) | 0.5# | 0.35 | 0.58# | 0.12 - 1.05 |
| Lab3_WWTP12 | Province 3 | Prov3_district 1 | 18000 | Medium | Fortnightly | 25 (8) | 0.4# | 0.07 | 0.41# | -0.12 - 0.94 |
| Lab3_WWTP13 | Province 3 | Prov3_district 1 | 48900 | Medium | Fortnightly | 32 (16) | 0.5# | 0.69 | 1.10 | 0.74 - 1.46 |
| **Total** |  |  |  |  |  | **154** (69) |  |  |  |  |
| **Lab 4 Lumegen** |  |  |  |  |  |  |  |  |  |  |
| Lab4_WWTP1 | Province 4 | Prov4_district 1 | 13200 | Medium | Fortnightly | 16 (11) | 0.83 | 0.68 | 2.39 | 1.38 - 3.40 |
| Lab4_WWTP2 | Province 4 | Prov4_district 1 | 645455 | Large | Fortnightly | 21 (18) | 0.2# | -0.02 | 0.55# | -0.92 - 2.01 |
| Lab4_WWTP3 | Province 4 | Prov4_district 1 | 20000 | Medium | Fortnightly | 12 (8) | 0.2# | -0.02 | 0.63# | -1.24 - 2.51 |
| Lab4_WWTP4 | Province 4 | Prov4_district 1 | 12500 | Medium | Fortnightly | 21 (15) | 0.5# | 0.19 | 1.47 | 0.04 - 2.90 |
| Lab4_WWTP5 | Province 4 | Prov4_district 1 | 17380 | Medium | Monthly | 10 (5) | 0.7# | 0.38 | 1.92# | -0.12 - 3.95 |
| Lab4_WWTP6 | Province 4 | Prov4_district 1 | 80000 | Medium | Fortnightly | 19 (15) | 0.6 | 0.20 | 1.74 | -0.05 - 3.53 |
| Lab4_WWTP7 | Province 5 | Prov5_district 1 | 426000 | Large | Fortnightly | 6 (6) | 0.4 | 0.25 | 1.16 | 0.32 - 2.00 |
| Lab4_WWTP8 | Province 6 | Prov6_district 1 | 40000 | Medium | Fortnightly | 38 (34) | 0.7 | 0.49 | 1.91 | 1.20 - 2.63 |
| Lab4_WWTP9 | Province 6 | Prov6_district 2 | 110159 | Medium | Fortnightly | 35 (28) | 0.6 | 0.27 | 1.23# | 0.46 - 2.00 |
| Lab4_WWTP10 | Province 7 | Prov7_district 1 | 248000 | Large | Fortnightly | 38 (34) | 0.5 | 0.21 | 1.27 | 0.29 - 2.25 |
| Lab4_WWTP11 | Province 8 | Prov8_district 1 | 250000 | Large | Fortnightly | 38 (33) | 0.6 | 0.33 | 1.44 | 0.65 - 2.23 |
| Lab4_WWTP12 | Province 8 | Prov8_district 2 | 185348 | Large | Fortnightly | 24 (24) | 0.4# | 0.03 | 0.31# | -0.72 - 1.34 |
| Lab4_WWTP13 | Province 8 | Prov8_district 3 | 509000 | Large | Fortnightly | 38 (33) | 0.7 | 0.37 | 1.94 | 1.01 - 2.88 |
| **Total** |  |  |  |  |  | **316 (264)** |  |  |  |  |
| **Lab 5 NICD** |  |  |  |  |  |  |  |  |  |  |
| Lab5_WWTP1 | Province 3 | Prov3_district 1 | 141000 | Medium | Weekly | 46 (24) | 0.3# | 0.03 | 0.22# | -0.06 - 0.51 |
| Lab5_WWTP2 | Province 3 | Prov3_district 1 | 112900 | Medium | Weekly | 45 (39) | 0.8 | 0.56 | 1.05 | 0.76 - 1.35 |
| Lab5_WWTP3 | Province 3 | Prov3_district 2 | 40000 | Medium | Weekly | 54 (48) | 0.8 | 0.52 | 0.93 | 0.28 - 1.6 |
| Lab5_WWTP4 | Province 3 | Prov3_district 2 | 100320 | Medium | Monthly | 9 (9) | 0.7 | 0.39 | 1.06 | 0.05 - 2.07 |
| Lab5_WWTP5 | Province 4 | Prov4_district 1 | 350000 | Large | Weekly | 45 (45) | 0.6 | 0.38 | 0.80 | 0.51 - 1.10 |
| Lab5_WWTP6 | Province 4 | Prov4_district 1 | 200000 | Large | Weekly | 46 (25) | 0.7 | 0.50 | 0.94 | 0.65 - 1.22 |
| Lab5_WWTP7 | Province 1 | Prov1_district 2 | 1200000 | Large | Monthly | 50 (50) | 0.6# | 0.08 | 0.30# | -0.20 - 0.79 |
| Lab5_WWTP8 | Province 1 | Prov1_district 1 | 350000 | Large | Weekly | 41 (41) | 0.7 | 0.44 | 0.82 | 0.51 - 1.13 |
| Lab5_WWTP9 | Province 1 | Prov1_district 2 | 500000 | Large | Weekly | 46 (44) | 0.7 | 0.40 | 0.66 | 0.42 - 0.90 |
| Lab5_WWTP10 | Province 1 | Prov1_district 3 | 6355 | Small | Weekly | 51 (51) | 0.7 | 0.55 | 0.87 | 0.65 - 1.09 |
| Lab5_WWTP11 | Province 1 | Prov1_district 3 | 100000 | Medium | Weekly | 49 (49) | 0.7 | 0.40 | 0.79 | 0.52 - 1.05 |
| Lab5_WWTP12 | Province 1 | Prov1_district 1 | 100000 | Medium | Monthly | 4 (44) | 0.4# | 0.00 | 1.00# | -3.34 - 5.35 |
| Lab5_WWTP13 | Province 1 | Prov1_district 1 | 350000 | Large | Monthly | 4 (4) | 1.0 | 0.74 | 0.67# | -2.62 - 3.96 |
| Lab5_WWTP14 | Province 2 | Prov2_district 1 | 316425 | Large | Fortnightly | 43 (42) | 0.7 | 0.38 | 0.67 | 0.42 - 0.92 |
| Lab5_WWTP15 | Province 7 | Prov7_district 2 | 10207 | Medium | Monthly | 15 (12) | 0.7 | 0.67 | 1.64 | 0.98 - 2.30 |
| Lab5_WWTP16 | Province 9 | Prov9_district 1 | 380000 | Large | Monthly | 11 (10) | 0.7 | 0.42 | 0.97 | 0.21 - 1.72 |
| Lab5_WWTP17 | Province 9 | Prov9_district 1 | 460000 | Large | Fortnightly | 30 (29) | 0.9 | 0.78 | 0.98 | 0.79 - 1.17 |
| Lab5_WWTP18 | Province 1 | Prov1_district 1 | 200000 | Large | Weekly | 45 (37) | 0.8 | 0.64 | 0.86 | 0.64 - 1.08 |
| **Total** |  |  |  |  |  | **634** (564) |  |  |  |  |
| **Lab 6** |  |  |  |  |  |  |  |  |  |  |
| Lab6_WWTP1 | Province 1 | Prov1_district 2 | 20000 | Medium | Weekly | 28 (25) | 0.4# | 0.16 | 0.61 | 0.13 - 1.10 |
| Lab6_WWTP2 | Province 1 | Prov1_district 2 | 100000 | Medium | Weekly | 30 (24) | 0.2# | 0.07 | 0.40# | -0.07 - 0.86 |
| Lab6_WWTP3 | Province 1 | Prov1_district 3 | 1368 | Small | Weekly | 50 (42) | 0.5 | 0.21 | 0.76 | 0.28 - 1.23 |
| Lab6_WWTP4 | Province 1 | Prov1_district 3 | 34051 | Medium | Weekly | 27 (14) | 0.5 | 0.33 | 0.54 | 0.17 - 0.98 |
| Lab6_WWTP5 | Province 1 | Prov1_district 3 | 2456 | Small | Weekly | 44 (40) | 0.2# | 0.09 | 0.45# | -0.00 - 0.91 |
| Lab6_WWTP6 | Province 1 | Prov1_district 3 | 866 | Small | Weekly | 15 (14) | 0.5 | 0.26 | 0.54 | 0.06 - 1.02 |
| Lab6_WWTP7 | Province 1 | Prov1_district 3 | 40000 | Medium | Weekly | 14 (12) | ## | ## | ## | ## |
| **Total** |  |  |  |  |  |  |  |  |  |  |
| **Lab 7** |  |  |  |  |  |  |  |  |  |  |
| Lab7_WWTP1 | Province 1 | Prov1_district 3 | 97417 | Medium | Fortnightly | 28 (21) | ## | ## | ## | ## |
| Lab7_WWTP2 | Province 1 | Prov1_district 3 | 40000 | Medium | Fortnightly | 23 (16) | ## | ## | ## | ## |
| Lab7_WWTP3 | Province 1 | Prov1_district 3 | 2500 | Small | Fortnightly | 19 (10) | ## | ## | ## | ## |
| **Total** |  |  |  |  |  | **70** (47) |  |  |  |  |
| **Lab 8** |  |  |  |  |  |  |  |  |  |  |
| Lab8_WWTP1 | Province 1 | Prov1_district 2 | 850000 | Large | Weekly | 20 (5) | 0.6 | 0.38 | 0.53 | 0.18 - 0.88 |
| Lab8_WWTP2 | Province 1 | Prov1_district 2 | 850000 | Large | Weekly | 42 (23) | 0.2# | -0.03 | 0.24# | -0.60 - 1.09 |
| Lab8_WWTP3 | Province 1 | Prov1_district 3 | 27343 | Medium | Fortnightly | 21 (21) | 0.4# | -0.02 | 0.33# | -0.05 - 1.13 |
| Lab8_WWTP4 | Province 1 | Prov1_district 3 | 87896 | Medium | Fortnightly | 37 (14) | -0.02# | -0.06# | 0.02# | -0.51 - 0.55 |
| Lab8_WWTP5 | Province 1 | Prov1_district 3 | 58431 | Medium | Monthly | 34 (14) | ## | ## | ## | ## |
| Lab8_WWTP6 | Province 1 | Prov1_district 3 | 9120 | Small | Fortnightly | 22 (2) | ## | ## | ## | ## |
| Lab8_WWTP7 | Province 1 | Prov1_district 3 | 9312 | Small | Weekly | 18 (5) | -0.2# | -0.05 | -0.24# | -1.08 - 0.60 |
| Lab8_WWTP8 | Province 1 | Prov1_district 1 | 100000 | Medium | Fortnightly | 21 (2) | ## | ## | ## | ## |
| Lab8_WWTP9 | Province 1 | Prov1_district 1 | 20000 | Medium | Fortnightly | 15 (15) | 0.6# | 0.21 | 0.48# | -0.12 - 1.07 |
| Lab8_WWTP10 | Province 1 | Prov1_district 1 | 20000 | Medium | Fortnightly | 33 (9) | 0.4# | 0.16 | 0.60 | 0.03 - 1.17 |
| Lab8_WWTP11 | Province 1 | Prov1_district 1 | 100000 | Medium | Weekly | 22 (7) | 0.7 | 0.49 | 0.76 | 0.54 - 0.98 |
| Lab8_WWTP12 | Province 1 | Prov1_district 1 | 100000 | Medium | Fortnightly | 21 (5) | -0.02# | -0.04 | -0.17# | -0.78 - 0.44 |
| Lab8_WWTP13 | Province 1 | Prov1_district 1 | 46789 | Medium | Fortnightly | 11 (0) | ## | ## | ## | ## |
| Lab8_WWTP14 | Province 1 | Prov1_district 1 | 10000 | Medium | Fortnightly | 23 (4) | 0.1# | -0.05 | 0.04# | -0.32 - 0.40 |
| Lab8_WWTP15 | Province 1 | Prov1_district 1 | 100000 | Medium | Fortnightly | 11 (1) | 0.4# | 0.2 | 0.49# | -0.02 - 0.99 |
| Lab8_WWTP16 | Province 1 | Prov1_district 1 | 10000 | Small | Monthly | 14 (14) | ## | ## | ## | ## |
| Lab8_WWTP17 | Province 1 | Prov1_district 4 | 6363 | Small | Fortnightly | 13 (13) | ## | ## | ## | ## |
| Lab8_WWTP18 | Province 1 | Prov1_district 4 | 11766 | Medium | Fortnightly | 12 (4) | ## | ## | ## | ## |
| Lab8_WWTP19 | Province 1 | Prov1_district 4 | 150 | Small | Fortnightly | 14 (0) | ## | ## | ## | ## |
| Lab8_WWTP20 | Province 2 | Prov2_district 1 | 30000 | Medium | Monthly | 17 (4) | ## | ## | ## | ## |
| Lab8_WWTP21 | Province 2 | Prov2_district 2 | 31518 | Medium | Monthly | 8 (3) | 0.3# | -0.02 | 0.69# | -1.32 - 2.69 |
| Lab8_WWTP22 | Province 5 | Prov5_district 1 | 426000 | Large | Monthly | 30 (9) | ## | ## | ## | ## |
| Lab8_WWTP23 | Province 5 | Prov5_district 1 | 12285 | Medium | Fortnightly | 9 (0) | ## | ## | ## | ## |
| Lab8_WWTP24 | Province 6 | Prov6_district 2 | 50000 | Medium | Fortnightly | 10 (2) | -0.3# | -0.01 | -0.17 | -0.91 - 0.58 |
| Lab8_WWTP25 | Province 8 | Prov8_district 3 | 509000 | Large | Fortnightly | 17 (2) | ## | ## | ## | ## |
| Lab8_WWTP26 | Province 9 | Prov9_district 1 | 1645000 | Large | Weekly | 25 (5) | ## | ## | ## | ## |
| Lab8_WWTP27 | Province 9 | Prov9_district 1 | 900000 | Large | Weekly | 25 (2) | ## | ## | ## | ## |
| Lab8_WWTP28 | Province 4 | Prov4_district 1 | 100000 | Medium | Fortnightly | 20 (2) | ## | ## | ## | ## |
| **Total** |  |  |  |  |  | **565** (153) |  |  |  |  |
| **Cumulative total** |  |  |  |  |  | **2328 (1536)** |  |  |  |  |

*Levels below the ‘level of quantification’ were reported as ‘less than the level of quantification in log copies/ml’

\# p≥0.005

\## absent or invalid results due to insufficient observations

** Population size and size category of plant not available
